# Point-of-care ultrasonography for risk stratification of non-critical suspected COVID-19 patients on admission (POCUSCO): a prospective binational study

**DOI:** 10.1101/2021.03.09.21253208

**Authors:** François Morin, Delphine Douillet, Jean François Hamel, Dominique Savary, Christophe Aubé, Karim Tazarourte, Kamélia Marouf, Florence Dupriez, Philipe Le Conte, Thomas Flament, Thomas Delomas, Mehdi Taalba, Nicolas Marjanovic, Francis Couturaud, Nicolas Peschanski, Thomas Boishardy, Jérémie Riou, Vincent Dubée, Pierre-Marie Roy

**Author notes:** **Corresponding author information** François Morin, MD, Univ Angers, University Hospital of Angers, Emergency Department, 4, rue Larrey, 49933 Angers Cedex 9, France.

## Abstract

**Background:** Lung point-of-care ultrasonography (L-POCUS) is highly effective in detecting pulmonary peripheral patterns and may allow early identification of patients who are likely to develop an acute respiratory distress syndrome (ARDS). We hypothesized that L-POCUS performed during the initial examination would identify non-severe COVID-19 patients with a high risk of getting worse.

**Methods:** POCUSCO was a prospective, multicenter study. Non-critical adult patients who were admitted to the emergency department (ED) for suspected or confirmed COVID-19 were included and had L-POCUS performed within 48 hours following admission. The severity of lung damage was assessed using the L-POCUS score based on 36 points for ARDS. The primary outcome was the rate of patients requiring intubation or who died within 14 days following inclusion.

**Results:** Among 296 participating patients, 8 (2.7%) had primary outcome. The area under the curve (AUC) of the receiver operating characteristic of L-POCUS was 0.80 [95%CI:0.60-0.94]. The score values which achieved a sensibility > 95% in defining low-risk patients and a specificity > 95% in defining high-risk patients were <1 and ≥16, respectively. The rate of patients with an unfavorable outcome was 0/95 (0%[95%CI:0-3.9]) for low-risk patients (score=0) versus 4/184 (2.17%[95%CI:0.8-5.5]) for intermediate-risk patients (score 1-15) and 4/17 (23.5%[95%CI:11.4-42.4]) for high-risk patients (score ≥16). In patients with confirmed COVID-19 (n=58), the AUC of L-POCUS was 0.97 [95%CI:0.92-1.00].

**Conclusions:** L-POCUS allows risk-stratification of patients with suspected or confirmed COVID-19. These results should be confirmed in a population with a higher risk of an unfavorable outcome.

**Trial registration number:** NCT04338100

## BACKGROUND

The COVID-19 pandemic has developed worldwide since its emergence in China in December 2019.[1–3] The majority of patients has a mild or uncomplicated course (81%) with minor symptoms such as headache, loss of smell, or cough. However, around 14% of patients develop respiratory symptoms and require hospitalization.[4] Median time from illness onset to dyspnea is 6 to 8 days and around 5% of the patients develop acute respiratory distress syndrome (ARDS), usually between Day 7 and Day 10.[4–6] The rapid progression of respiratory failure soon after the onset of dyspnea is a striking feature of COVID-19.[7],[8] There is an urgent need for reliable tools which can identify patients who are likely to get worse and develop ARDS early on.

Pulmonary computed tomography (CT-scan) appears to be very sensitive (97%) and quite specific for diagnosis of COVID-19 in patients with a clinical suspicion, provided that it is not performed within the first 4 days after symptom onset.[9,10] COVID-19 manifests itself on CT-scans as bilateral, subpleural, ground-glass opacities with air bronchograms, and ill-defined margins.[11] Those patterns can precede the positivity of the Reverse Transcriptase-Polymerase Chain Reaction (RT-PCR) for SARS-CoV-2.[12,13]

Lung point-of-care ultrasonography (L-POCUS) is a simple, non-invasive, non-irradiating, inexpensive imaging tool that is available at the bedside and used more and more by emergency physicians in their everyday clinical practice. L-POCUS seems to be better than chest X-ray in detecting pneumonia and may be an alternative to the CT-scan as a screening and prognostic tool.[14] Indeed, L-POCUS is highly effective in detecting peripheral patterns and pleural abnormalities, and seems appropriate for triaging COVID-19 patients.[15] A recent review highlights its potential value in decision making for triage or follow-up.[16] Many physicians have placed their hopes in this device, as shown by the number of publications reporting personal experiences or case reports but few prospective studies have been carried out on this topic, and, to our knowledge, no robust data have been yet provided on the prognostic value of L-POCUS in COVID-19 patients.

The aim of this study is to determine the performance of L-POCUS at the time of ED admission in identifying, among patients with confirmed or highly suspected COVID-19, those who are at high-risk of adverse outcomes such as respiratory failure or death.

## METHODS

### Study design and participants

The point-of-care ultrasonography for risk stratification of COVID-19 patients’ study (POCUSCO) was a non-interventional, prospective, multicenter study that was conducted in 11 participating hospitals in France and Belgium.

Patients were enrolled if they met all of the following criteria: (1) adult patients (≥ 18 years old); (2) typical COVID-19 symptoms and at least one of the three following features: i) positive SARS-CoV-2 RT-PCR, ii) typical CT-scan lesions, iii) COVID-19 is the main diagnostic hypothesis by the in-charge physician; (3) no requirement for respiratory support and/or other intensive care, and not subject to a limitation of care; (4) membership of a social security scheme.

Patients for whom the follow-up at Day 14 was impossible or who had a condition making lung ultrasonography impossible (body mass index > 35 kg/m^2^, history of pneumonectomy) were excluded.

The initial evaluation was carried out by the physician-in-charge and patients were treated as standard.[17] All participating patients underwent L-POCUS and a score reflecting the intensity and the extension of lung involvement was determined.[18] This score was previously developed for ARDS (see below).[18,19] For patients who were subsequently hospitalized, a second L-POCUS was performed on Day 5 ± 3 under the same conditions as the first one, whenever it was possible.

Patients were followed up by phone at Day 14 and their clinical status recorded according to the Ordinal Scale for Clinical Improvement for COVID-19 from the World Health Organization (WHO-OSCI) (Table 1).[20]

**Table 1.** Ordinal Scale for Clinical Improvement (OSCI) of the World Health Organization (WHO)

| Patient state | Descriptor | Score |
| --- | --- | --- |
| Uninfected | No clinical or virological evidence of infection | 0 |
| Ambulatory | No limitation of activities | 1 |
|  | Limitation of activities | 2 |
| Hospitalized Mild Disease | Hospitalized, no oxygen therapy | 3 |
|  | Oxygen by mask or nasal prongs | 4 |
| Hospitalized Severe Disease | Non-invasive ventilation or high-flow oxygen | 5 |
|  | Intubation and mechanical ventilation | 6 |
|  | Ventilation + additional organ support: pressors, renal replacement therapy, ECMO... | 7 |
| Dead | Death | 8 |

### Objectives and outcomes

The main objective was to assess the ability of L-POCUS to identify COVID-19 patients with a high-risk of an unfavorable outcome. The primary endpoint was the development of severe COVID-19 within the 14 days after ED admission defined as a stage of the WHO-OSCI ≥ 6. This stage relates to a severe inpatient requiring intubation and invasive ventilation (stage 6), and/or additional organ support (stage 7) or who died whatever the cause (stage 8). The ability of L-POCUS to predict the primary outcome occurrence was evaluated by the area under the curve (AUC) of the receiver operating characteristic (ROC) curve and its 95% confidence interval (95%CI). A sensitivity analysis was performed with the 14-day all-cause mortality rate as the outcome.

The secondary objectives were:

1. To determine the threshold values of L-POCUS to perform risk stratification in three groups of patients: low-risk patients, intermediate-risk patients, and high-risk patients.
2. To assess the impact of adding the result of POCUS evaluation to two risk-stratification clinical scores: the quick Sequential Organ Failure Assessment (qSOFA) and the CRB-65.[21,22]
3. To assess the impact of the knowledge and experience of the operator level (novice, confirmed or expert) on the L-POCUS performance.

We performed a subgroup analysis in patients for whom the diagnosis of COVID-19 was initially or subsequently confirmed by a positive RT-PCR for SARS-CoV-2.

### Lung point-of-care ultrasonography

L-POCUS was performed with ultrasound scanners using low frequency (2-5 MHz) transductors, convex or small linear type probes. The Bedside Lung Ultrasound in an Emergency (BLUE)-Protocol was applied to patients in erect or semi-recumbent positions depending on dyspnea severity (Figure 1).[19] Each chest wall was divided by the anterior and posterior axillary lines into anterior, lateral, or posterior regions. All intercostal spaces of the upper and lower parts of these regions were examined, resulting in a total of 12 areas of investigation. Each area was examined for at least one complete respiratory cycle. Four ultrasound aeration patterns were defined and scored 0 to 3, allowing calculation of the L-POCUS score, theoretically ranging from 0 to 36 (Figure 1).[18,23] Considering biological risk of infection, special protective precautions were taken to protect the operator and other patients as recommended.[24]

**Figure 1.**
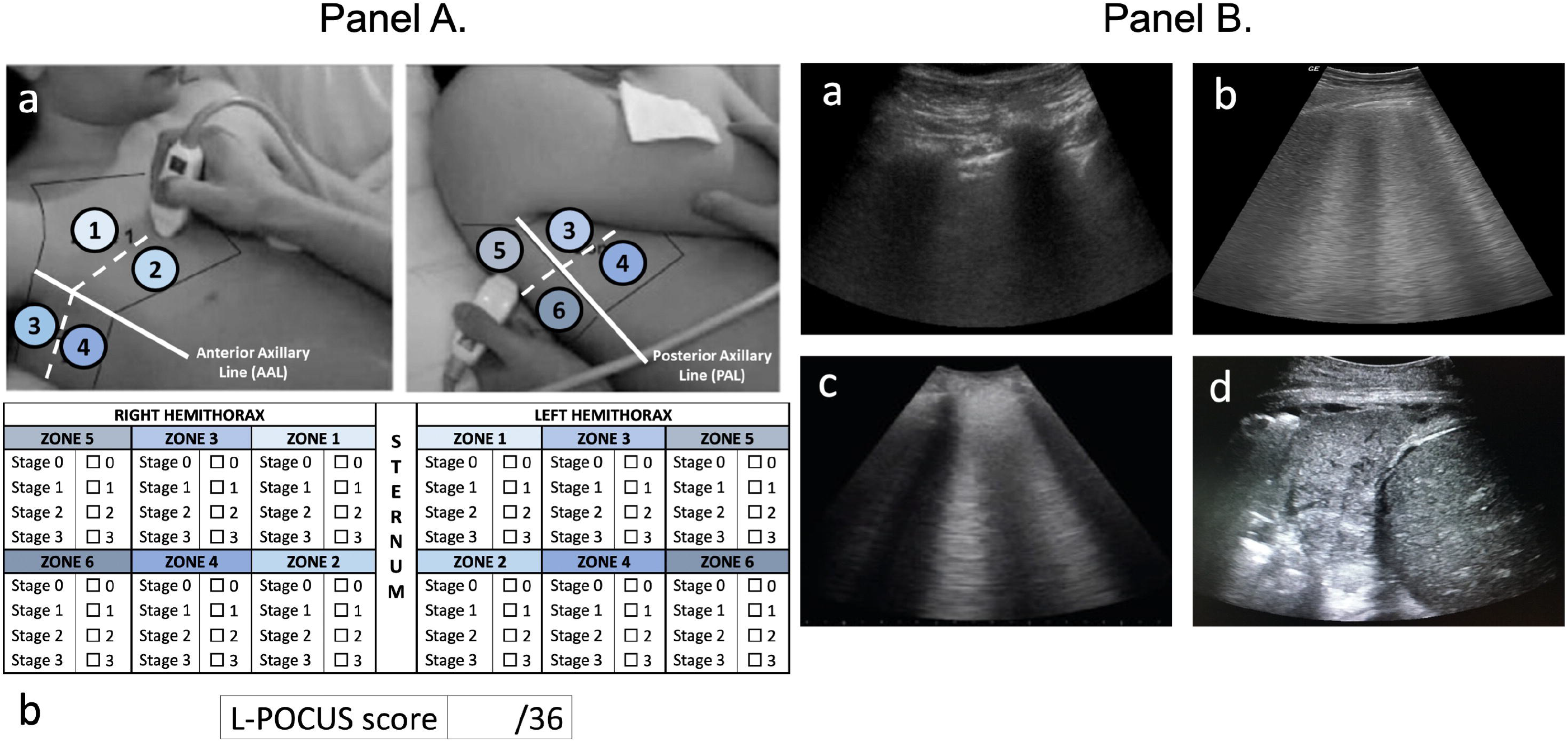
**Panel A. Lung Point Of Care Ultrasonography Method (L-POCUS)** a. Twelve chest areas of investigation following BLUE-PLUS Protocol: *zone 1*: upper anterior chest wall; *zone 2*: lower anterior chest wall; *zone 3*: upper lateral chest wall; *zone 4*: lower lateral chest wall; *zone 5*: upper posterolateral chest wall; *zone 6*: lower posterolateral chest wall b. L-POCUS score grid: Each zone was examined to establish which of four ultrasound parenchymal aeration stages it exhibited, and points are assigned to them according to their severity. *<u>Stage 0 or normal aeration (0 point)</u>*: Lung sliding sign associated with respiratory movement of less than 3 B lines; *<u>Stage 1 or moderate loss of lung aeration (1 point)</u>*: a clear number of multiple visible B-lines with horizontal spacing between adjacent B lines□≤□7 mm (B1 lines); *<u>Stage 2 or severe loss of lung aeration (2 points)</u>*: multiple B lines fused together that were difficult to count with horizontal spacing between adjacent B lines□≤□3 mm, including “white lung”; and *<u>Stage 3 or pulmonary consolidation (3 points)</u>*: hyperechoic lung tissue, accompanied by dynamic air bronchogram **Panel B. Examples of four ultrasound aeration stages** a. *Stage 0 or normal aeration b. Stage 1 or moderate loss of lung aeration; c. Stage 2 or severe loss of lung aeration; d. Stage 3 or pulmonary consolidation*.

### Ethics and financials

This study was conducted in accordance with the Declaration of Helsinki, as amended. The protocol was approved by the Ethics Committee CPP Sud-Ouest et Outre-Mer II for France (No. 2020-A00782-37 / 2-20-025 id7566) and the Ethics Committee of the Cliniques Universitaires Saint-Luc for Belgium (No. 2020/14AVR/223). Written informed consent was obtained from all patients. The study was funded by a grant from the French Ministry of Health (PHRC-I, April 2020, COVID19_A_001). This study adheres to STROBE guidelines, and all its details have been verified before submitting the manuscript.[25]

### Statistical analyses

Continuous variables were expressed as mean and standard deviation values. Categorical variables were described using numbers, percentages and their 95% confidence intervals (95%CI). The AUCs and their 95% confidence interval were determined by the .632 bootstrap method. For the primary outcome, we determined in advance that the L-POCUS prognostic value would be considered as clinically relevant with a good level of evidence if the lower bound of the 95%CI of the AUC was equal to or greater than 0.7. To perform risk stratification in three groups of patients with a low, intermediate, or high-risk of an unfavorable outcome, two thresholds were calculated. The first maximized specificity with a sensitivity greater than or equal to 95% and the second maximized sensitivity with a specificity greater than or equal to 95%. For these threshold values, sensitivity, specificity, predictive values and likelihood ratios were assessed. To study the impact of adding the results of the L-POCUS evaluation to several risk stratification clinical rules for pulmonary infection or sepsis (qSOFA and CRB65), AUCs were compared with or without their components with a DeLong test. For this purpose, we attributed 0, 1, or 2 points in the L-POCUS result as low, moderate or high risk according to the predefined threshold values and assessed the AUC of the risk-stratification rules with and without adding the L-POCUS result value. Assuming a rate of death or tracheal intubation requirement of 10%, and expecting an AUC of 0.8, the number of patients required to achieve a lower limit of the 95%CI, more than 0.7, was estimated as 286. Taking into consideration that 5% of patients were not followed up or could not be evaluated, the sample size was defined as 300 patients. Missing data were not imputed. A descriptive analysis of missing data was performed and compared to the available data to assess a potential bias. All statistical analyzes were performed using STATA, version 14.2; StataCorp; College Station, TX.

## RESULTS

### Characteristics of study population

A total of 307 patients with suspected or confirmed SARS-CoV-2 infection were enrolled in this study. Among them, 2 were subsequently excluded and 9 were could not be followed up (2.93%) leaving 296 patients for the main analyses (Figure 2). The mean age of the overall population was 57 years (± 20.8), and 146 (47.6%) were men (Table 2). The more common symptoms of COVID-19 were dyspnea (n=230 [74.9%]) cough (n=193 [62.9%]), abnormal thoracic auscultation (n=149 [48.5%]) and chest pain (n=125 [40.7%]).

**Table 2:** Demographic and clinical characteristics of participating patients

|  | All patients<br>(N = 307) |
| --- | --- |
| **Epidemiological characteristics** |  |
| Age (years), mean ± SD | 56.94 ± 20.76 |
| Gender, N (%) |  |
| Male | 146 (47.6%) |
| Female | 161 (52.4%) |
| **Comorbidities, N (%)** |  |
| Neurovascular diseases | 24 (7.8%) |
| COPD | 26 (8.5%) |
| Asthma | 46 (15.0%) |
| Hypertension | 104 (33.9%) |
| Diabetes | 37 (12.0%) |
| Active neoplasia | 18 (5.9%) |
| Chronic renal failure | 19 (6.2%) |
| Hepatic insufficiency | 6 (1.9%) |
| Chronic heart failure | 26 (8.5%) |
| **Clinical characteristics, N (%)** |  |
| Confusion or GCS < 15 | 10 (3.3%) |
| Cough | 193 (62.9%) |
| Anosmia/ ageusia/ dysgeusia | 50 (16.3%) |
| Dyspnea | 230 (74.9%) |
| Rhinorrhea | 52 (16.9%) |
| Diarrhea | 61 (19.9%) |
| Abnormal pulmonary auscultation | 149 (48.5%) |
| Chest pain | 125 (40.7%) |
| **Vital parameters** |  |
| Heart rate (bpm), mean ± SD | 90.67 ± 18.30 |
| SBP (mmHg), mean ± SD | 136.43 ± 22.48 |
| Temperature (°C), mean ± SD | 37.16 ± 1.00 |
| SpO2 (%), mean ± SD | 96.55 ± 3.09 |
| Respiratory rate (cpm), mean ± SD | 21.96 ± 6.08 |
| Oxygenotherapy, N (%) | 85 (27.7%) |

*RT-PCR: reverse transcriptase polymerase chain reaction; COPD: chronic obstructive pulmonary disease; GCS: Glasgow coma scale; SBP: systolic blood pressure; SpO2: pulse-oxymetry; qSOFA score: quick sepsis related organ failure assessment score; C(U)RB-65 score: pneumonia scores based on confusion/(urea)/respiratory rate/blood pressure/age ≥ 65*

**Figure 2.**
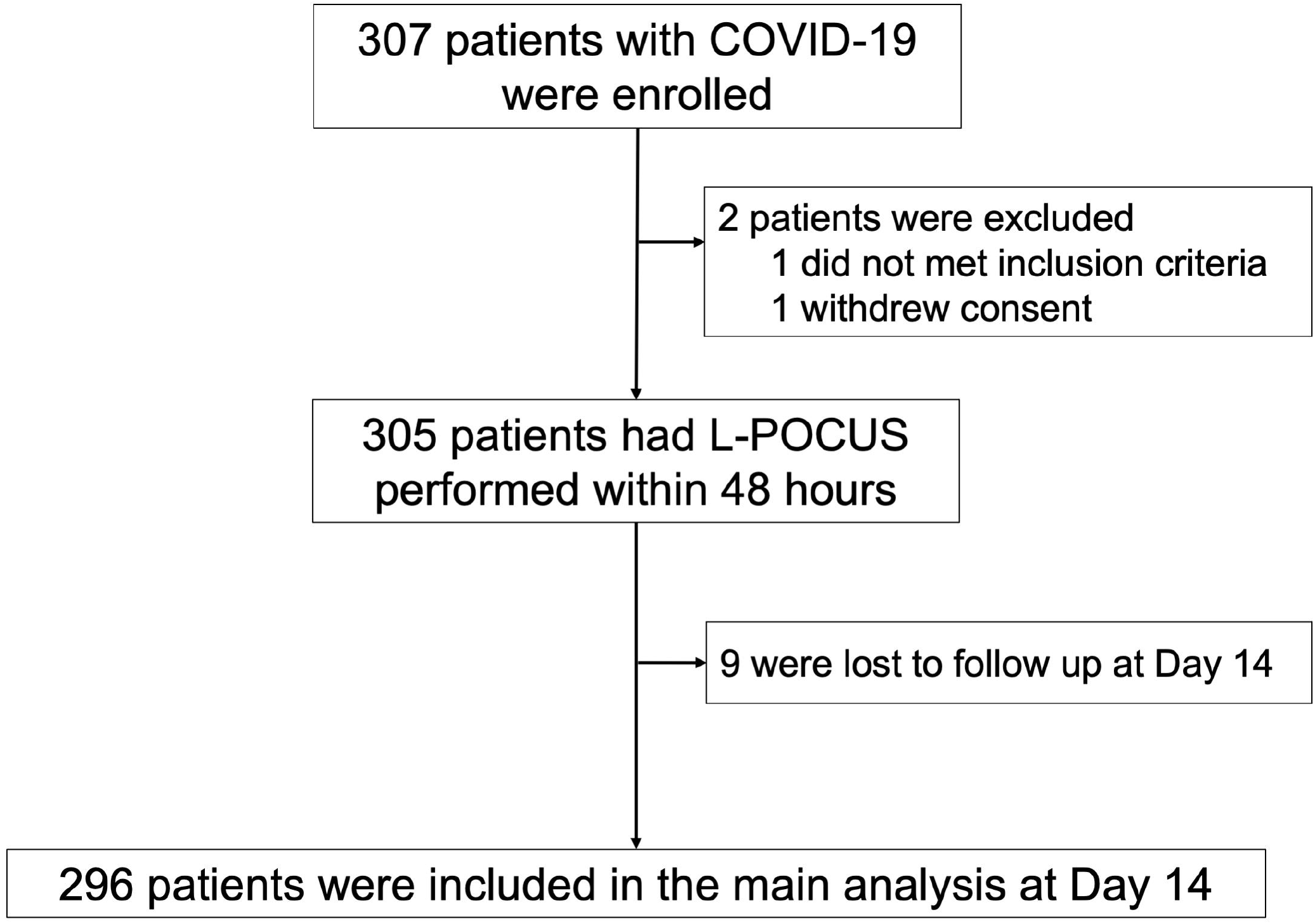
Study flow chart. COVID-19: Coronavirus disease 2019; L-POCUS: lung point of care ultrasonography; OSCI: ordinal scale for clinical improvement

The L-POCUS was performed by an emergency physician considered an expert, an advanced technician and a novice in 32.2% (n=99), 44.3% (n=133) and 24.4% (n=75) of cases respectively. A CT-scan was performed on 170 patients (55.4%) and 40 patients (22.9%) had a second L-POCUS at Day 5 ± 3.

### Outcomes in the overall population

The results of the L-POCUS are outlined in Figure 3. At Day 14, among 296 analyzable patients, the main outcome occurred in 8 (2,7%) patients (seven were dead and one patient had required intubation and invasive ventilation). The AUC of L-POCUS was 0.80 (95%CI: 0.60-0.94) (Figure 4). The lower value of the 95% CI did not achieve the predefined value of 0.7 necessary to consider the performance of L-POCUS as clinically relevant. In the sensitivity analysis with the 14-day all-cause mortality rate as an outcome, the AUC of L-POCUS was 0.83 (95%CI: 0.66-1).

**Figure 3.**
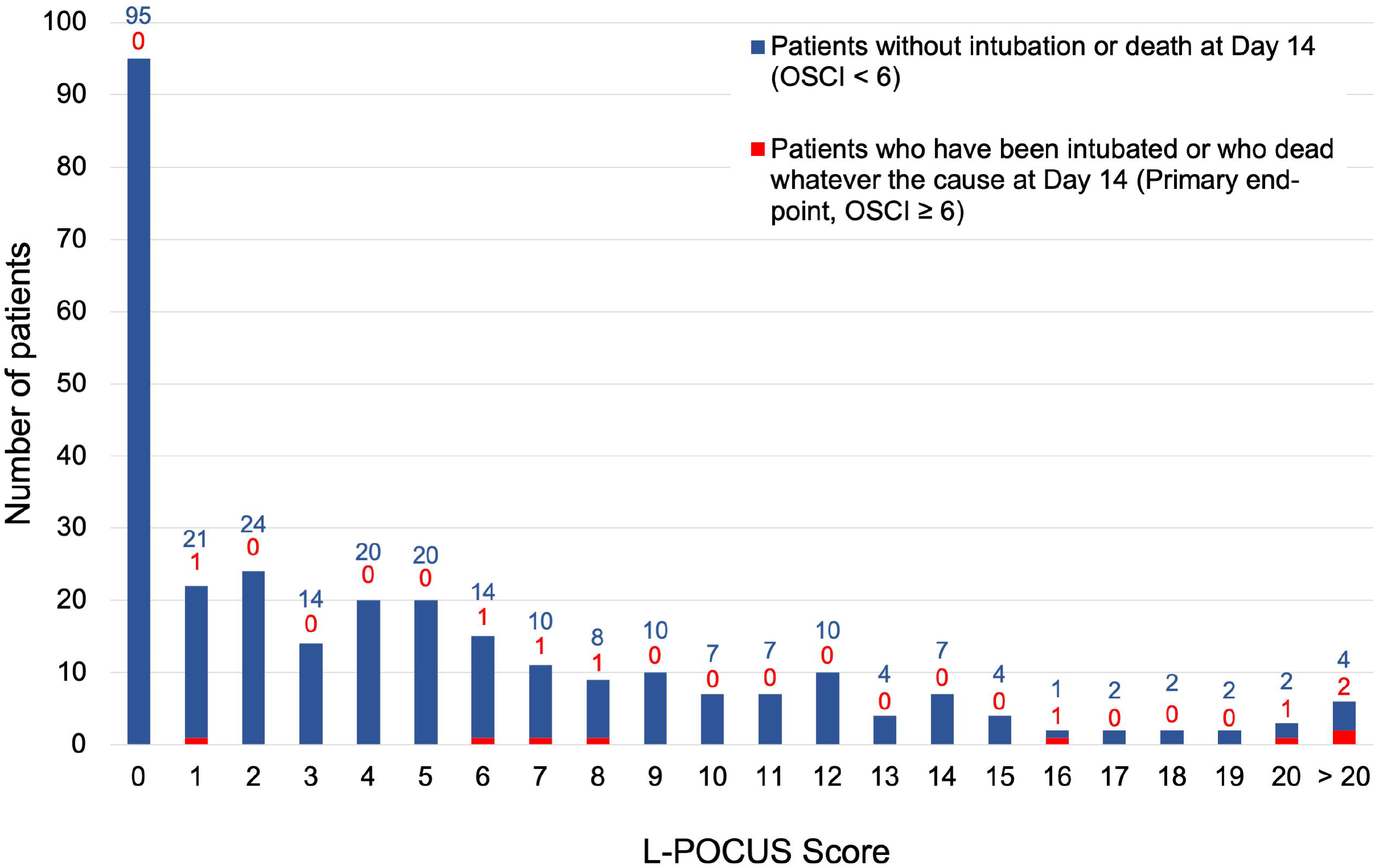
Distribution of L-POCUS score according to Ordinal Scale for Clinical Improvement at Day 14. Ordinal Scale for Clinical Improvement (OSCI) < 6 (blue); OSCI ≥ 6 (red)

**Figure 4.**
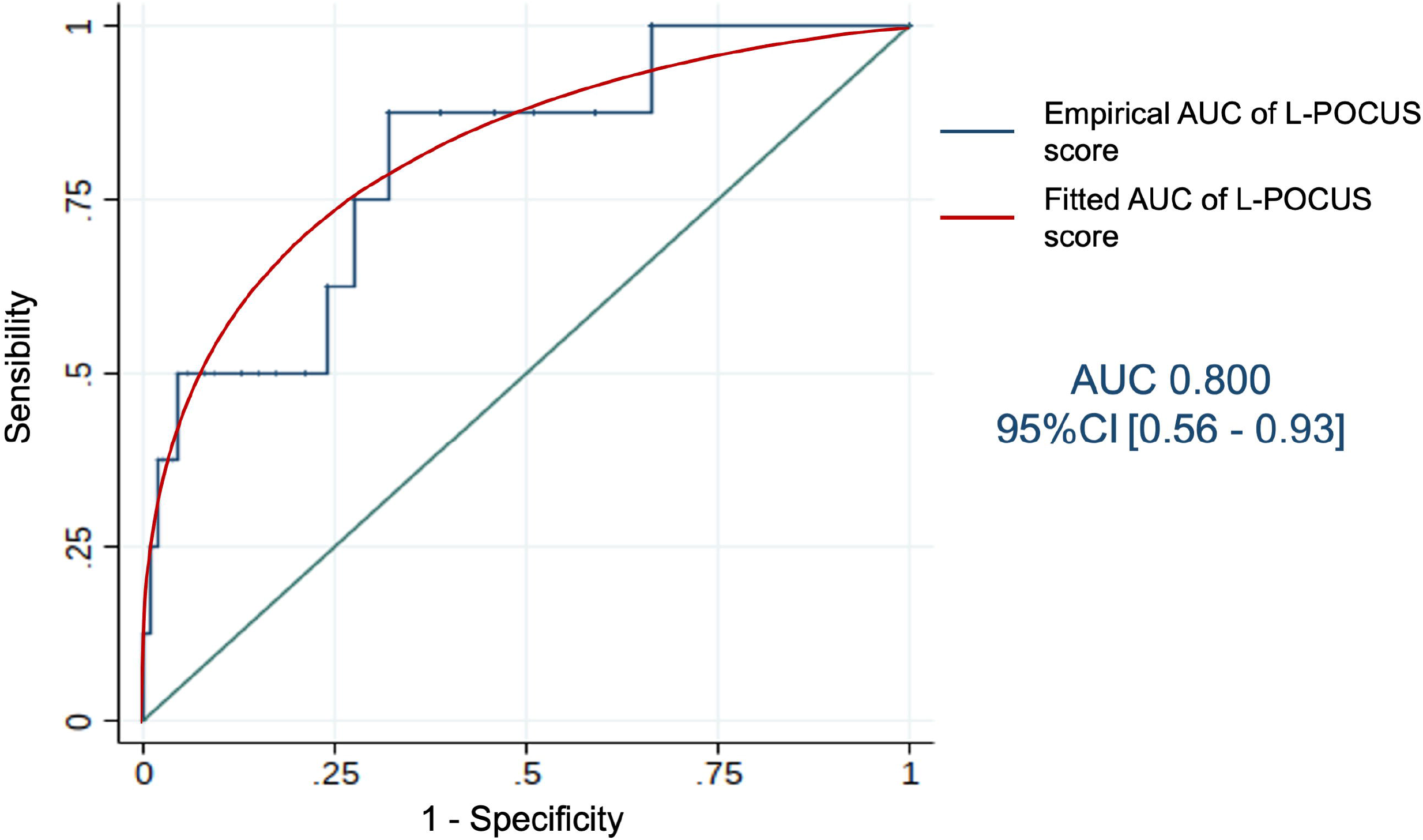
L-POCUS prognostic performance. Receiver operating characteristic (ROC)) curve of prognostic performance of global L-POCUS with its area-under-the-curve (AUC) and its 95% confidence interval (95%CI)

The AUC slightly increased according to the experience of the POCUS operator without significant difference: 0.86 (95%CI: 0.70-0.99), 0.82 (95%CI: 0.34-1) and 0.68 (95%CI: 0.56-0.78), for experts, confirmed or novices, respectively.

The highest L-POCUS with a sensitivity of at least 95% was 0 point and the lowest value with a specificity of at least 95% was 16 points. Using these cutoffs, 95 patients (32.1%) had a low-risk (score =0) and none of them had an unfavorable outcome at Day 14 (0% [95%CI: 0.0-3.9]; Sensitivity 100% [95%CI: 63.1-100.0]; Specificity 33.0% [95%CI: 27.6-38.7]; Positive likelihood ratio (LR+) 1.49 [95%CI: 1.4-1.6]; Negative likelihood ratio (LR-) 0; Positive predictive value (PPV) 3.9% [95%CI: 3.7-4.3]; Negative predictive value (NPV) 100%). 184 patients (62.4%) had intermediate-risk (score 1 to 15) and, among them, 4 (2.17% [95%CI: 0.8-5.5]) had an unfavorable outcome (Positive likelihood ratio (LR+) 0.8 [95%CI: 0.5-1.3]; Negative likelihood ratio (LR-) 1.33 [95%CI: 0.8-2.1]). Finally, 17 patients (5.7%) had a high-risk (score ≥ 16) and, among them, 4 (23.5%) had an unfavorable outcome at Day 14 (23.5% [95%CI: 11.4-42.4]; Sensitivity 50% [95%CI: 15.7-84.3]; Specificity 95.5% [95%CI: 92.4-97.6]; LR+ 11.1 [95%CI: 4.6-26.5]; LR-0.5 [95%CI: 0.3-1.1]; PPV 23.5% [95%CI: 11.4-42.4]; NPV 98.6% [95%CI: 97.2-99.3]).

The AUCs of the risk prediction clinical rules qSOFA and CRB65 with and without addition of the L-POCUS score were 0.75 [95%CI: 0.56 to 0.94] and 0.52 [95%CI: 0.32 to 0.71], and 0.82 [95%CI: 0.68 to 0.99] and 0.72 [95%CI: 0.49 to 0.95], respectively.

### Patients with Positive SARS-CoV-2

Among 240 patients tested (78.2%), 58 (24.2%) had a positive RT-PCR for SARS-CoV2. At Day 14, 4 patients with confirmed COVID-19 were dead (4/58, 6.9%). The AUC of L-POCUS was 0.97 (95%CI: 0.92 – 1.00). Using the two thresholds defined in the overall cohort, L-POCUS determined 6 patients (10.5%) with low-risk and none of them had an unfavorable outcome at Day 14 (0% [95%CI: 0 to 21.5]; Sensitivity 100% [95%CI: 39.8-100]; Specificity 11.3% [95%CI: 4.3-23.0]). 43 patients (75.4%) presented an intermediate-risk and none of them had an unfavorable outcome (0% [95%CI: 0-8.2]). Among 8 patients (14.0%) presenting a high-risk, 4 had an unfavorable outcome (50.0% [95%CI: 23.7-76.3-Sensitivity 50% [95%CI: 15.7-84.3]; Specificity 92.0% [95%CI: 80.8-97.8].

## DISCUSSION

In our prospective POCUSCO study of non-severe patients with confirmed or suspected COVID-19, L-POCUS has had good results in predicting the occurrence of death or requirement for invasive ventilation within the 14 days following ED admission and it appears to be a promising tool for risk stratification. However, because of a lower-than-expected rate of patients with an unfavorable outcome, the confidence intervals of our estimates are wide with an upper value of the AUC not achieving the predefined value of 0.7 to consider the L-POCUS prognostic value as clinically relevant with a good level of evidence.

Based on its performance in diagnosing pneumonia and ARDS, L-POCUS ought to be a useful diagnostic and risk stratification tool in the initial assessment of suspected COVID-19 patients.[14,26,27] It is currently considered an alternative to physical examination for suspected COVID-19 patients in the emergency department.[27] However, this position is mainly based on expert opinion and few trials have been published. Moreover, most of them are monocentric studies assessing the correlation of L-POCUS with chest CT scans in detecting lung abnormalities suggestive of COVID-19 and/or its value in diagnosing patients with suspected COVID-19. Globally, they suggest a high sensitivity at around 90% but with a low specificity at around 25%, depending on disease prevalence.[28,29] Those estimates are greater than the first RT-PCR value.[29] L-POCUS would provide an effective estimate of the extent of the pulmonary histological damage.[30]

To our knowledge, only one previous study assessed the performance of L-POCUS in identifying patients with suspected or confirmed COVID-19 at risk of deteriorating. Indeed, Bonadia et al. conducted a prospective study in 41 COVID-19 patients and showed the higher the rate, the higher the in-hospital mortality and need for intensive care admission.[31] Our results therefore provide further important data regarding prognostication and triage with L-POCUS.

Ultrasonography including L-POCUS was questioned for its lack of reproducibility, being dependent on the examiner. To avoid this pitfall, standardized procedures have been proposed.[32] We used a revised BLUE protocol previously validated in patients with ARDS.[18] Based on the screening of twelve chest areas and on weighting with four aeration patterns, this score is quick and easy to achieve, which is particularly relevant in the Emergency Department and in the context of the strain on health resources.[18] Aeration patterns are easy to recognize and the use in our study of pocket cards with the L-POCUS score (Figure 1) make it easy for most emergency physicians to apply. It is important to note that in previous studies, L-POCUS were performed by experienced emergency physicians, all certified for lung ultrasound.[33,34] In our trial, nearly a quarter of the exams were performed by novices physicians without any significant difference in terms of the AUC of L-POCUS from the exams performed by experts. Indeed, a short training with 25 supervised L-POCUS helps novices acquire skills in L-POCUS.[35]

With an AUC of 0.80, the global performance of L-POCUS is good in our overall population and similar results were observed by considering only patients who died as an outcome. They are even better in the subgroup of patients with positive RT-PCR for SARS-CoV2, the lower limit of the AUC being higher than 0.9. These results are particularly relevant in the current context of organized mass screening. Unlike at the start of the pandemic, most of the patients consulting the ED have the results of testing or could benefit from a quick test for COVID-19. Moreover, recent data suggest that “thickening of the pleural lining, may be an important pattern for L-POCUS assessment of the prognosis of COVID-19 patients.[36] The inclusion of this criterion in a revised version of the score in a future study may improve the risk-stratification performance of L-POCUS for COVID-19 patients.

In terms of implementing L-POCUS as a triaging tool in every day clinical practice, we aimed to stratify the result into three risk categories. None of the 95 patients who were determined to be low-risk (L-POCUS score = 0) suffered significant deterioration and home treatment may be suitable for these patients, if they lack comorbidities or a living condition which precludes this option. It is important to note that only one patient with a L-POCUS score < 6 had an unfavorable outcome within the 14 days following ED admission. This patient was not positive with SARS-CoV-2 and died from pulmonary malignancy. On the other hand, 4 of 17 patients determined as high risk (score ≥ 16) died. All of them had a positive RT-PCR SARS-CoV-2 and died from COVID-19. Nevertheless, these results must be considered carefully before using L-POCUS in the early triage of COVID-19 patients, at least as a standalone tool.

In our trial the prognostic performance of the qSOFA and CRB-65 were low but the addition of the L-POCUS to these clinical rules slightly improved their performance in terms of the AUC: +0.23 for qSOFA and +0.1 for CRB-65. These results are in line with the study of Bar et al showing that a model combining the qSOFA and 4 ultrasound findings has good value as a diagnostic tool (AUC: 0.82 [95%CI: 0.75 – 0.90]) of.[37] The best result was obtained with CRB-65 + L-POCUS with an AUC of 0.82 [95%CI: 0.68 to 0.99]. However, the lower limit of the 95%CI did not achieve the prespecified level of 0.7.

To our knowledge, POCUSCO is the largest multicentric, prospective study evaluating L-POCUS to risk-stratify COVID-19 patients. Nevertheless, it has some limitations, one of the more important being the low primary endpoint rate. On the basis of the first cohorts of COVID-19 inpatients, we considered a rate of mortality or invasive ventilation requirement of 10%, at the time the protocol was written.[38] It was actually 7% in confirmed COVID-19 patients and only 2.4% in our overall cohort. Several factors may explain this discrepancy: differences in the completeness of testing and case identification, variable thresholds for hospitalization and Intensive Care Unit admission, and improvement in patients’ care.[39] Moreover, only a quarter of participating patients had a positive RT-PCR for SARS-CoV-2, the other patients may have had a minor form of COVID-19 or another less severe disease. Nevertheless, our results are in line with the 1.4% mortality rate and 2.3% rate of patients who underwent invasive mechanical ventilation in the cohort of Guan et al.[4] Finally, in the absence of a derivation model, it is not methodologically justified to assess the calibration of L-POCUS.[18] Another study must be carried out to validate our results on an independent cohort.

## INTERPRETATION

L-POCUS allows risk-stratification of suspected or confirmed COVID-19 patients. Using a 36-points score initially defined for ARDS, L-POCUS determined patients with an exact score of to be low-risk and patients with a score ≥ 16 to be high-risk of death or of requiring invasive ventilation. Further studies are needed to confirm these results and to determine whether a global multimodal model, integrating L-POCUS score value and other considerations, would enable the risk stratification of COVID-19 patients more precisely than the L-POCUS score alone.

## Data Availability

All data referred to in the manuscript are be available after obtaining the agreement of the corresponding author.

## Notes

### Competing Interest Statement

The authors have declared no competing interest.

### Clinical Trial

NCT04338100

### Clinical Protocols

https://bmjopen.bmj.com/content/11/2/e041118

### Funding Statement

This study was carried out with a grant provided by the French Ministry of Health (PHRC-I, April 2020, COVID19_A_001). The sponsor had no role in the design of the study, the collection and analysis of the data, or the preparation of the manuscript. Apart from this grant, the authors declare no support from any organization for the submitted work.

### Author Declarations

This study was conducted in accordance with the Declaration of Helsinki, as amended. The protocol was approved by the Ethics Committee CPP Sud-Ouest et Outre-Mer II for France (No. 2020-A00782-37 / 2-20-025 id7566) and the Ethics Committee of the Cliniques Universitaires Saint-Luc for Belgium (No. 2020/14AVR/223).

## REFERENCES

1 Ye Q, Wang B, Mao J, et al. Epidemiological analysis of COVIDL19 and practical experience from China. J Med Virol Published Online First: 10 April 2020. doi:10.1002/jmv.25813

2 Novel Coronavirus Pneumonia Emergency Response Epidemiology Team. [The epidemiological characteristics of an outbreak of 2019 novel coronavirus diseases (COVID-19) in China]. Zhonghua Liu Xing Bing Xue Za Zhi 2020;41:145–51. doi:10.3760/cma.j.issn.0254-6450.2020.02.003

3 Wu Z, McGoogan JM. Characteristics of and Important Lessons From the Coronavirus Disease 2019 (COVID-19) Outbreak in China: Summary of a Report of 72L314 Cases From the Chinese Center for Disease Control and Prevention. JAMA Published Online First: 24 February 2020. doi:10.1001/jama.2020.2648

4 Guan W-J, Ni Z-Y, Hu Y, et al. Clinical Characteristics of Coronavirus Disease 2019 in China. N Engl J Med Published Online First: 28 February 2020. doi:10.1056/NEJMoa2002032

5 Zhou F, Yu T, Du R, et al. Clinical course and risk factors for mortality of adult inpatients with COVID-19 in Wuhan, China: a retrospective cohort study. Lancet Published Online First: 11 March 2020. doi:10.1016/S0140-6736(20)30566-3

6 Rodriguez-Morales AJ, Cardona-Ospina JA, Gutiérrez-Ocampo E, et al. Clinical, laboratory and imaging features of COVID-19: A systematic review and meta-analysis. Travel Med Infect Dis 2020;:101623. doi:10.1016/j.tmaid.2020.101623

7 Berlin DA, Gulick RM, Martinez FJ. Severe Covid-19. New England Journal of Medicine 2020;0:null. doi:10.1056/NEJMcp2009575

8 Tobin MJ, Laghi F, Jubran A. Why COVID-19 Silent Hypoxemia is Baffling to Physicians. Am J Respir Crit Care Med Published Online First: 15 June 2020. doi:10.1164/rccm.202006-2157CP

9 Ding X, Xu J, Zhou J, et al. Chest CT findings of COVID-19 pneumonia by duration of symptoms. Eur J Radiol 2020;127:109009. doi:10.1016/j.ejrad.2020.109009

10 Bernheim A, Mei X, Huang M, et al. Chest CT Findings in Coronavirus Disease-19 (COVID-19): Relationship to Duration of Infection. Radiology 2020;295:200463. doi:10.1148/radiol.2020200463

11 Shi H, Han X, Jiang N, et al. Radiological findings from 81 patients with COVID-19 pneumonia in Wuhan, China: a descriptive study. The Lancet Infectious Diseases 2020;20:425–34. doi:10.1016/S1473-3099(20)30086-4

12 Ai T, Yang Z, Hou H, et al. Correlation of Chest CT and RT-PCR Testing in Coronavirus Disease 2019 (COVID-19) in China: A Report of 1014 Cases. Radiology 2020;:200642. doi:10.1148/radiol.2020200642

13 Xie X, Zhong Z, Zhao W, et al. Chest CT for Typical 2019-nCoV Pneumonia: Relationship to Negative RT-PCR Testing. Radiology 2020;:200343. doi:10.1148/radiol.2020200343

14 Schenck EJ, Rajwani K. Ultrasound in the diagnosis and management of pneumonia. Curr Opin Infect Dis 2016;29:223–8. doi:10.1097/QCO.0000000000000247

15 Mohamed MFH, Al-Shokri S, Yousaf Z, et al. Frequency of Abnormalities Detected by Point-of-Care Lung Ultrasound in Symptomatic COVID-19 Patients: Systematic Review and Meta-Analysis. Am J Trop Med Hyg 2020;103:815–21. doi:10.4269/ajtmh.20-0371

16 Nouvenne A, Zani MD, Milanese G, et al. Lung Ultrasound in COVID-19 Pneumonia: Correlations with Chest CT on Hospital admission. RES 2020;99:617–24. doi:10.1159/000509223

17 World Health Organisation. Clinical management of COVID-19. 2020. https://www.who.int/publications-detail-redirect/clinical-management-of-covid-19 (xaccessed 24 Jun 2020).

18 Zhao Z, Jiang L, Xi X, et al. Prognostic value of extravascular lung water assessed with lung ultrasound score by chest sonography in patients with acute respiratory distress syndrome. BMC Pulm Med 2015;15:98. doi:10.1186/s12890-015-0091-2

19 Lichtenstein DA, Mezière GA. Relevance of lung ultrasound in the diagnosis of acute respiratory failure: the BLUE protocol. Chest 2008;134:117–25. doi:10.1378/chest.07-2800

20 World Health Organisation. WHO. R&D Blueprint - novel Coronavirus - COVID-19 Therapeutic Trial Synopsis. 2020.https://www.who.int/blueprint/priority-diseases/key-action/COVID-19_Treatment_Trial_Design_Master_Protocol_synopsis_Final_18022020.pdf

21 Ferreira FL, Bota DP, Bross A, et al. Serial evaluation of the SOFA score to predict outcome in critically ill patients. JAMA 2001;286:1754–8. doi:10.1001/jama.286.14.1754

22 Lim WS, Eerden MM van der, Laing R, et al. Defining community acquired pneumonia severity on presentation to hospital: an international derivation and validation study. Thorax 2003;58:377–82. doi:10.1136/thorax.58.5.377

23 Lichtenstein D. Échographie pulmonaire en réanimation et aux urgences. Réanimation 2008;17:722– 30. doi:10.1016/j.reaurg.2008.09.008

24 Skulstad H, Cosyns B, Popescu BA, et al. COVID-19 pandemic and cardiac imaging: EACVI recommendations on precautions, indications, prioritization, and protection for patients and healthcare personnel. Eur Heart J Cardiovasc Imaging 2020;21:592–8. doi:10.1093/ehjci/jeaa072

25 Cuschieri S. The STROBE guidelines. Saudi J Anaesth 2019;13:S31–4. doi:10.4103/sja.SJA_543_18

26 Soldati G, Smargiassi A, Inchingolo R, et al. Is There a Role for Lung Ultrasound During the COVID-19 Pandemic? Journal of Ultrasound in Medicine;n/a. doi:10.1002/jum.15284

27 Buonsenso D, Pata D, Chiaretti A. COVID-19 outbreak: less stethoscope, more ultrasound. The Lancet Respiratory Medicine 2020;0. doi:10.1016/S2213-2600(20)30120-X

28 Colombi D, Petrini M, Maffi G, et al. Comparison of admission chest computed tomography and lung ultrasound performance for diagnosis of COVID-19 pneumonia in populations with different disease prevalence. Eur J Radiol 2020;133:109344. doi:10.1016/j.ejrad.2020.109344

29 Pivetta E, Goffi A, Tizzani M, et al. Lung Ultrasonography for the Diagnosis of SARS-CoV-2 Pneumonia in the Emergency Department. Ann Emerg Med Published Online First: 13 October 2020. doi:10.1016/j.annemergmed.2020.10.008

30 de Almeida Monteiro RA, Duarte-Neto AN, Ferraz da Silva LF, et al. Ultrasound assessment of pulmonary fibroproliferative changes in severe COVID-19: a quantitative correlation study with histopathological findings. Intensive Care Med Published Online First: 3 January 2021. doi:10.1007/s00134-020-06328-4

31 Bonadia N, Carnicelli A, Piano A, et al. Lung Ultrasound findings are associated with mortality and need of intensive care admission in COVID-19 patients evaluated in the Emergency Department. Ultrasound in Medicine and Biology 2020;0. doi:10.1016/j.ultrasmedbio.2020.07.005

32 Soldati G, Smargiassi A, Inchingolo R, et al. Proposal for International Standardization of the Use of Lung Ultrasound for Patients With COVID-19: A Simple, Quantitative, Reproducible Method. J Ultrasound Med 2020;39:1413–9. doi:10.1002/jum.15285

33 Tung-Chen Y, Gracia MM de, Díez-Tascón A, et al. Correlation between Chest Computed Tomography and Lung Ultrasonography in Patients with Coronavirus Disease 2019 (COVID-19). Ultrasound in Medicine and Biology 2020;46:2918–26. doi:10.1016/j.ultrasmedbio.2020.07.003

34 Benchoufi M, Bokobza J, Chauvin AA, et al. Lung injury in patients with or suspected COVID-19L: a comparison between lung ultrasound and chest CT-scanner severity assessments, an observational study. medRxiv 2020;:2020.04.24.20069633. doi:10.1101/2020.04.24.20069633

35 Rouby J-J, Arbelot C, Gao Y, et al. Training for Lung Ultrasound Score Measurement in Critically Ill Patients. Am J Respir Crit Care Med 2018;198:398–401. doi:10.1164/rccm.201802-0227LE

36 Zhang Y, Xue H, Wang M, et al. Lung Ultrasound Findings in Patients With Coronavirus Disease (COVID-19). American Journal of Roentgenology 2020;:1–5. doi:10.2214/AJR.20.23513

37 Bar S, Lecourtois A, Diouf M, et al. The association of lung ultrasound images with COVID-19 infection in an emergency room cohort. Anaesthesia;n/a. doi:10.1111/anae.15175

38 Zhou F, Yu T, Du R, et al. Clinical course and risk factors for mortality of adult inpatients with COVID-19 in Wuhan, China: a retrospective cohort study. The Lancet 2020;0. doi:10.1016/S0140-6736(20)30566-3

39 Wiersinga WJ, Rhodes A, Cheng AC, et al. Pathophysiology, Transmission, Diagnosis, and Treatment of Coronavirus Disease 2019 (COVID-19): A Review. JAMA 2020;324:782–93. doi:10.1001/jama.2020.12839

